## Supplementary Materials for "Characterizing SARS-CoV-2 neutralization profiles after bivalent boosting using antigenic cartography"

**Supplementary Figures**

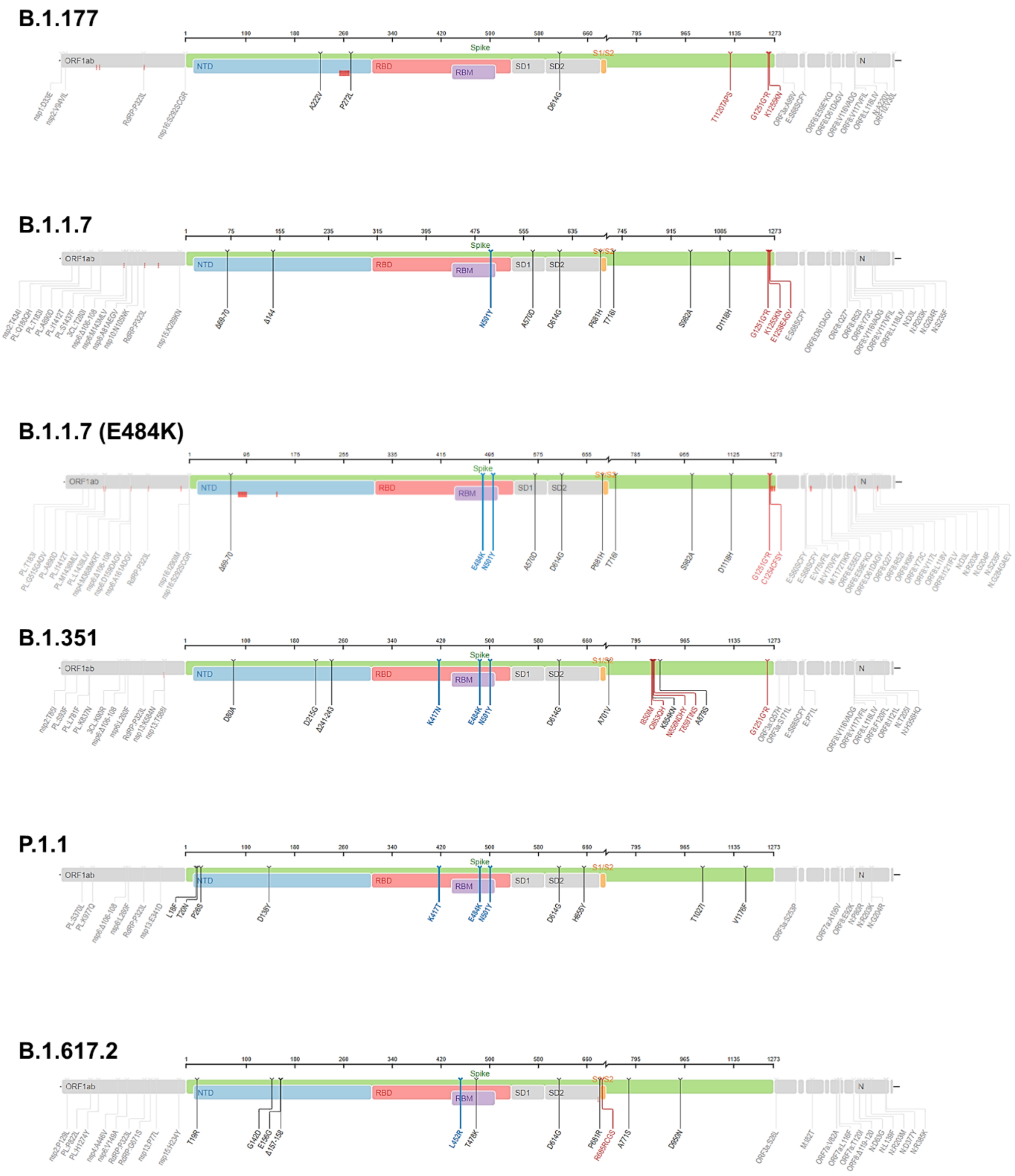


**Supplementary Figure 1. Spike mutations of pre-omicron virus isolates.** Graphics depicting spike mutations in pre-omicron variants (see Supplementary Table 2 for GISAID ID of isolated virus stocks) relative to Wuhan-1 were generated using https://covdb.stanford.edu/sierra/sars2/by-sequences/.

**BA.1**

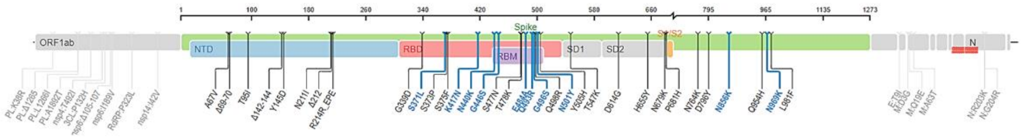


**Supplementary Figure 2. Spike mutations of BA.1 omicron virus isolate.** Graphics depicting spike mutations in BA.1 omicron variant (see Supplementary Table 2 for GISAID ID of isolated virus stock) relative to Wuhan-1 were generated using https://covdb.stanford.edu/sierra/sars2/by-sequences/.

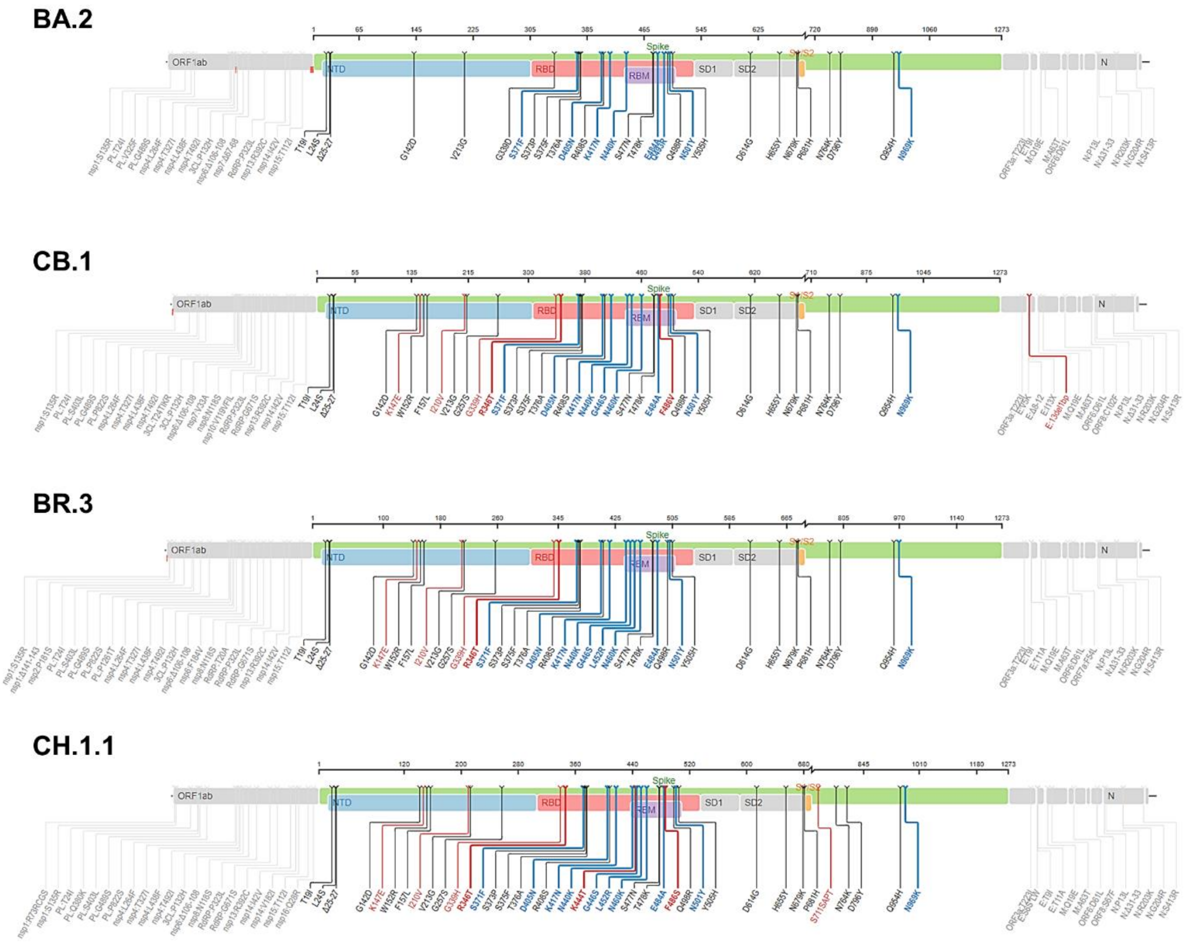


**Supplementary Figure 3. Spike mutations of BA.2 omicron virus isolates.** Graphics depicting spike mutations in pre-omicron variants (see Supplementary Table 2 for GISAID ID of isolated virus stocks) relative to Wuhan-1 were generated using https://covdb.stanford.edu/sierra/sars2/by-sequences/.

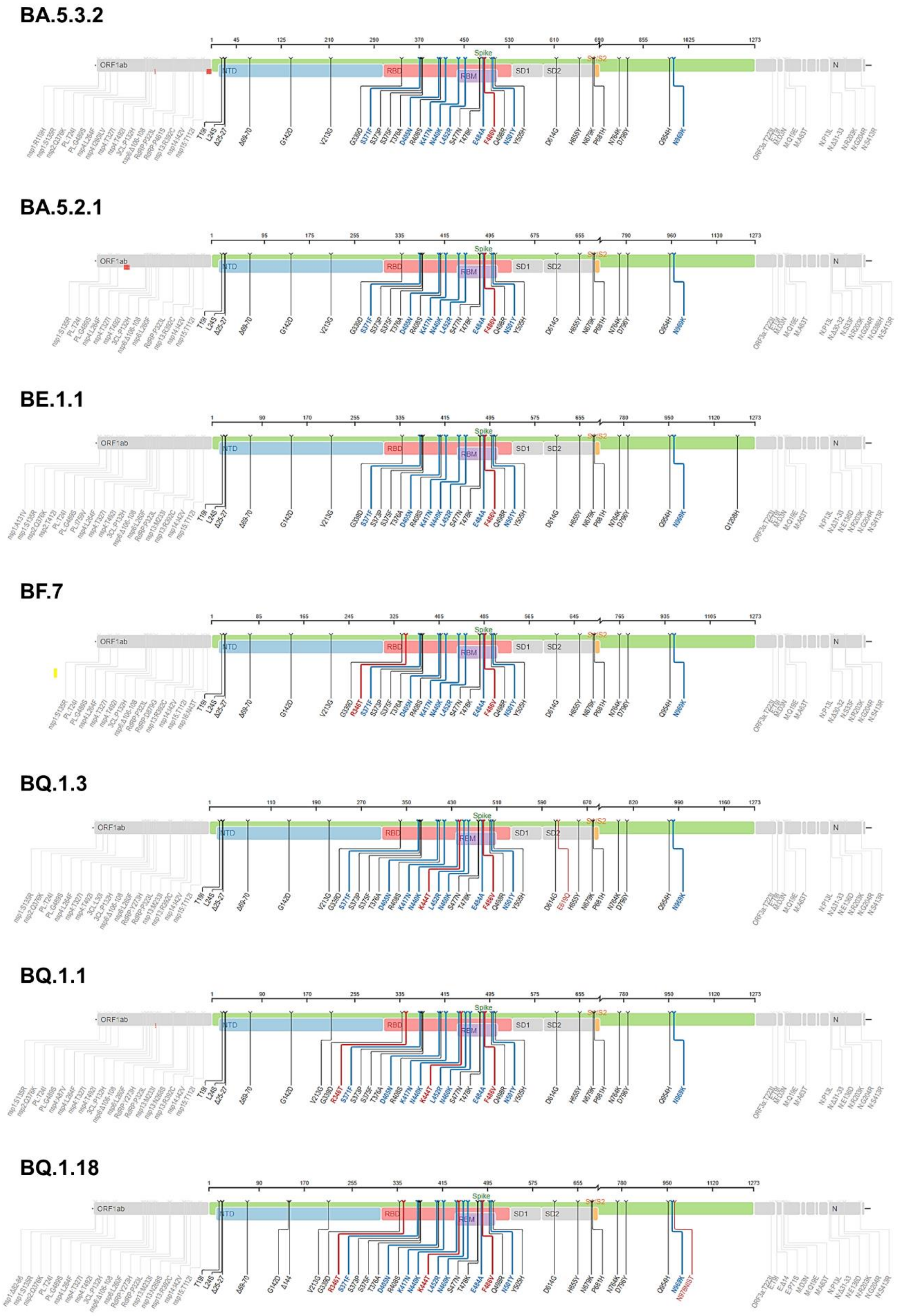


**Supplementary Figure 4. Spike mutations of BA.5 omicron virus isolates.** Graphics depicting spike mutations in pre-omicron variants (see Supplementary Table 2 for GISAID ID of isolated virus stocks) relative to Wuhan-1 were generated using https://covdb.stanford.edu/sierra/sars2/by-sequences/.

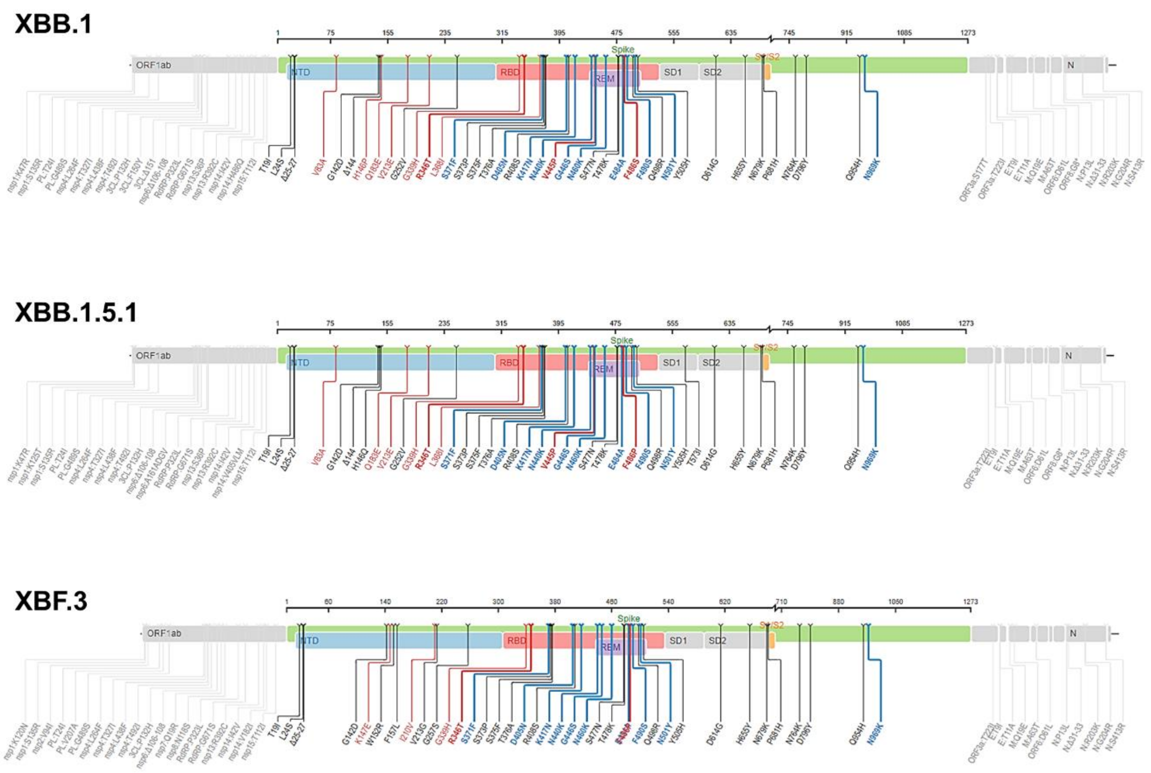


**Supplementary Figure 5. Spike mutations of recombinant omicron virus isolates.** Graphics depicting spike mutations in recombinant variants (see Supplementary Table 2 for GISAID ID of isolated virus stocks) relative to Wuhan-1 were generated using https://covdb.stanford.edu/sierra/sars2/by-sequences/.

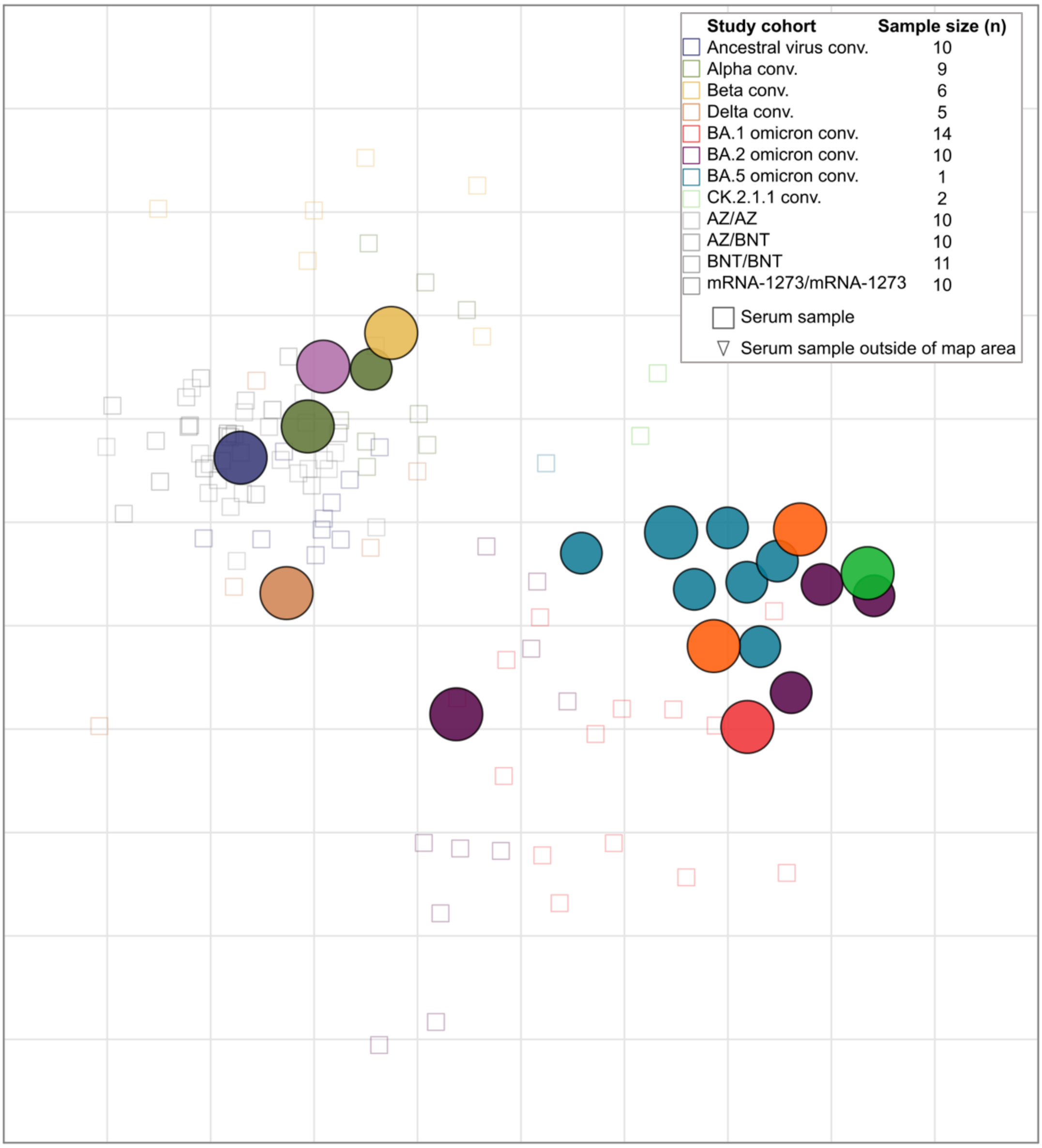


**Supplementary Figure 6. Non-zoomed in version of the antigenic map.** Virus variants are shown as colored circles, sera as open squares with the color corresponding to the infecting variant, vaccine sera are shown in grey tones. A smaller circle denotes variants with additional substitutions from the root variant (alpha+E484K, BA.5 and BA.2.75 sub-lineages). The x- and y-axis represent antigenic distances with one grid square corresponding to one two-fold serum dilution of the neutralization titer. The map orientation within x- and y-axis is free as only relative distances can be inferred. Only single variant exposure sera and double vaccination sera have been used for construction of the map. See Supplementary Table 3 for numbers of sera and virus variants used for calculation of the map.

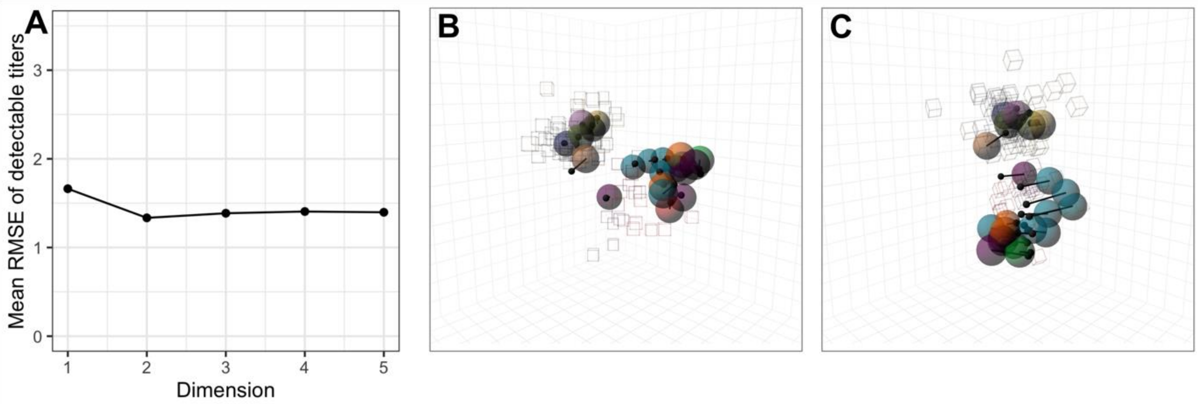


**Supplementary Figure 7. Map dimensionality. A** Dimensionality test: RMSE between map and measured titers for detectable titers in 1 to 5 dimensions. Per dimension, 100 map replicates were constructed from 90% of measured titers with 1000 optimizations per replicate. The titers of the remaining 10% were predicted in each run and the RMSE calculated by comparing the predicted to the measured titers on the $\log_2$ scale. **B, C** Side and front view of the map optimized in 3 dimensions with arrows pointing to the variants' position in the 2D map.

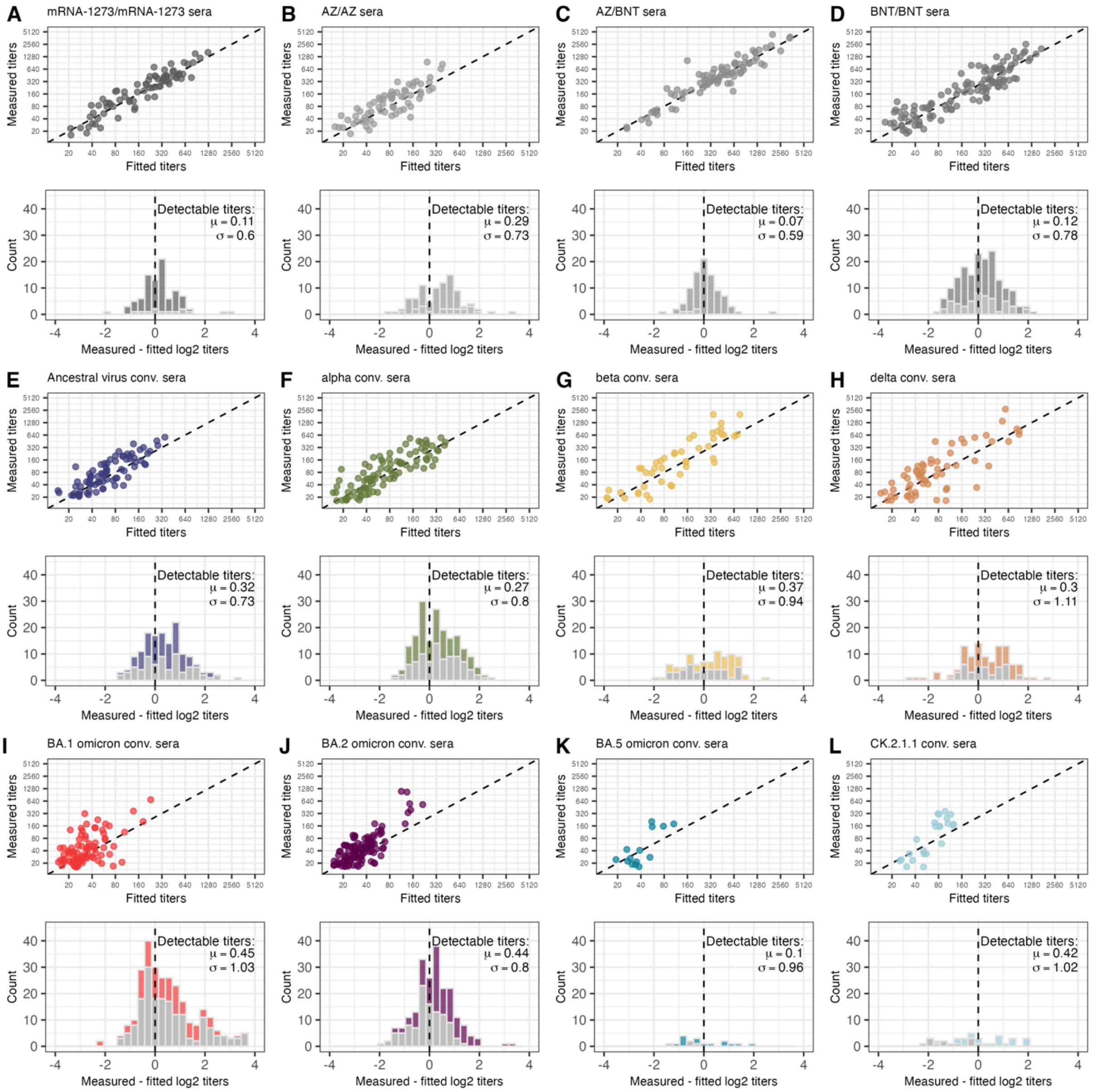


**Supplementary Figure 8. Goodness of map fit per serum group.** The top panels show the correlation of detectable measured and fitted titers in the 2D map with P.1.1 reactivity adjustment. Measured P.1.1 titers were reduced by one two-fold to match the reactivity adjustment in the map. Map distances were converted into $\log_2$ titers by subtracting the Euclidean distance for each serum-antigen pair from the maximum $\log_2$ titer of the specific serum. The bottom panels show the residuals of measured against fitted titers on the $\log_2$ scale, light grey marks pairs with the measured titer below the assay detection threshold. The mean and mean-centered standard deviation of differences between fitted and detectable measured titers are given in the legend of each bottom row panel. This was done for the serum groups used to construct the map: **A** mRNA-1273/mRNA-1273, **B** ChAdOx-S1/ChAdOx-S1, **C** ChAdOx-S1/BNT162b2, **D** BNT162b2/BNT162b2, **E** Ancestral virus conv., **F** alpha/alpha+E484K conv., **G** beta conv., **H** delta conv., **I** BA.1 omicron conv., **J** BA.2 omicron conv., **K** BA.5 omicron conv., **L** CK.2.1.1 conv..

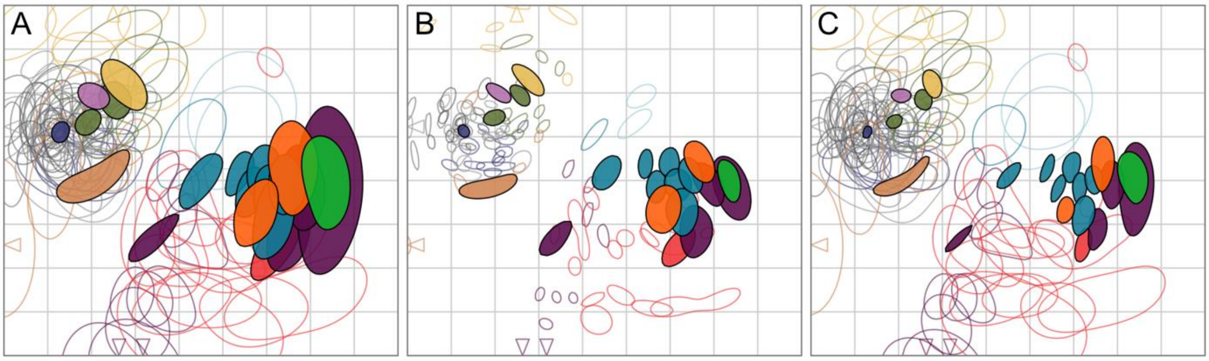


**Supplementary Figure 9. Assessing map robustness by bootstrapping.** 500 bootstrap repeats were performed with 1000 optimizations per repeat. In each repeat, different weights are assigned to each part of the titer table. The weights are drawn randomly from a Dirichlet distribution. Different weights were added to **A** titers and antigen reactivity, **B** only titers, and **C** only antigen reactivity. The colored regions mark 68% (one standard deviation) of the positional variation for each variant (filled shapes) and sera (open shapes). The colors correspond to the colors used in Figure X.

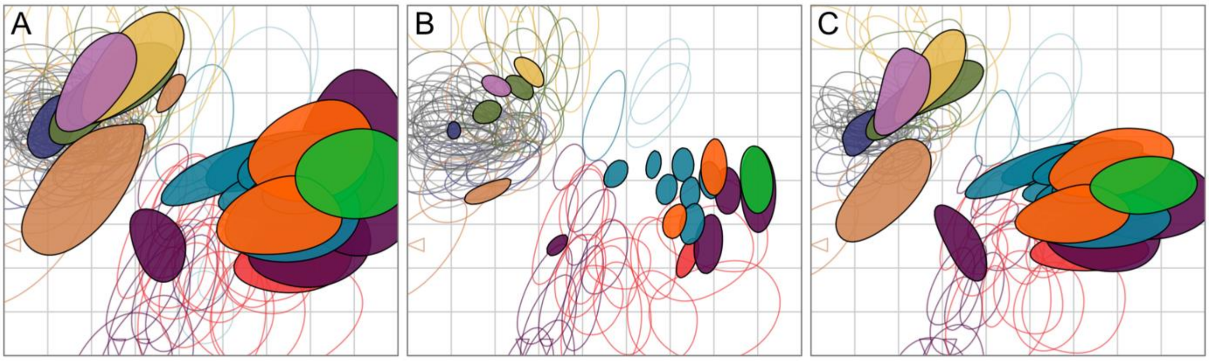


**Supplementary Figure 10. Assessing map robustness to measurement uncertainty by bootstrapping.** 500 bootstrap repeats were performed with 1000 optimizations per repeat. Normally distributed measurement noise with a standard deviation of 0.7 was added to **A** titers and antigen reactivity, **B** only titers, and **C** only antigen reactivity. The colored regions mark 68% (one standard deviation) of the positional variation for each variant (filled shapes) and sera (open shapes). The colors correspond to the colors used in Figure X.

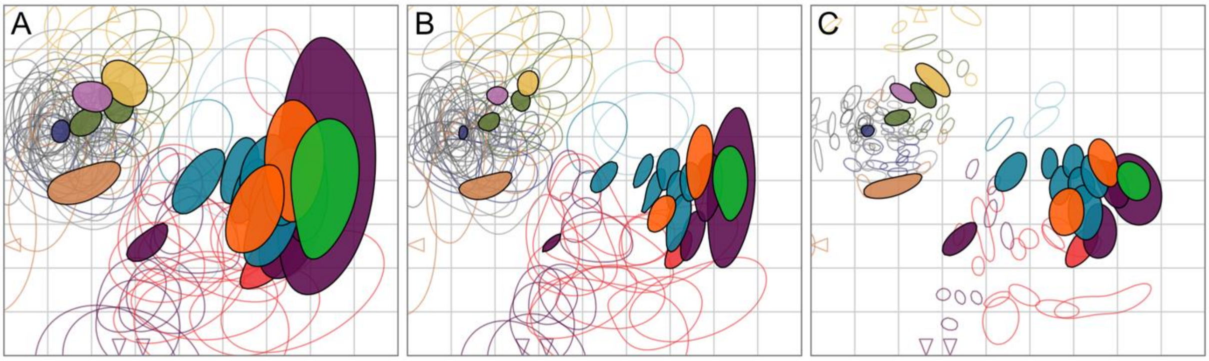


**Supplementary Figure 11. Assessing map robustness to the exclusion of measurements by bootstrapping.** 500 bootstrap repeats were performed with 1000 optimizations per repeat. For each repeat, a random subset of titer measurements was taken with replacement. The bootstrapping was performed on **A** variants and sera, **B** only variants, and **C** only sera. The colored regions mark 68% (one standard deviation) of the positional variation for each variant (filled shapes) and sera (open shapes). The colors correspond to the colors used in Figure X.

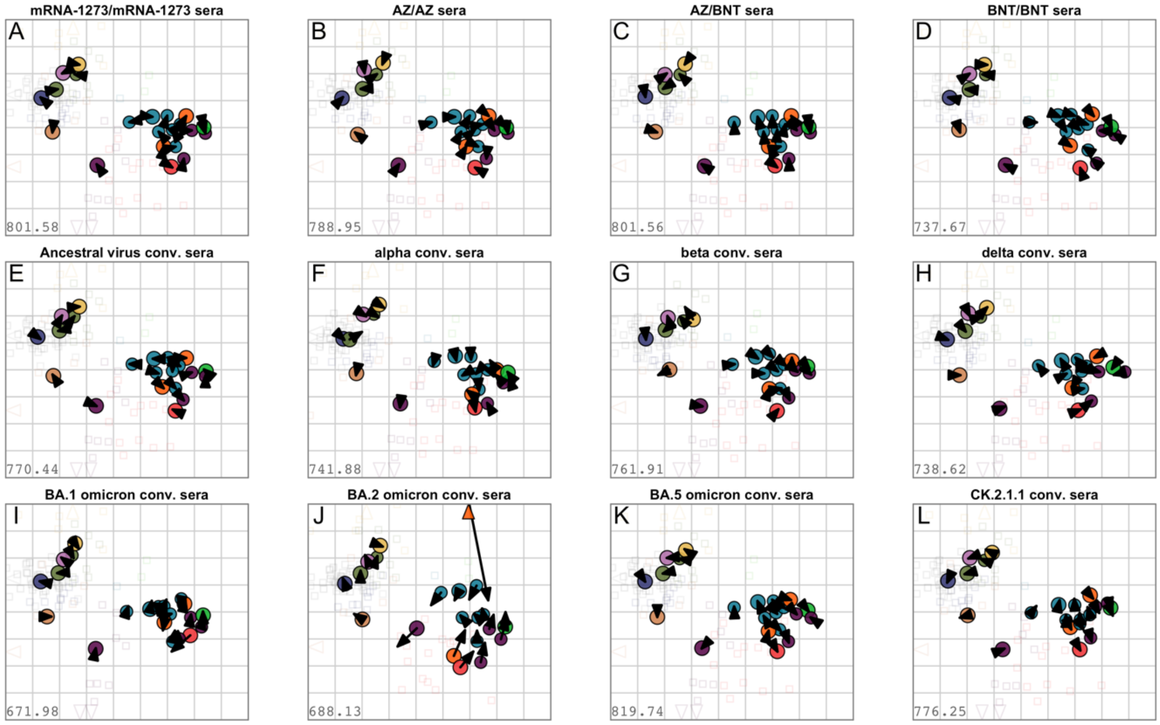


**Supplementary Figure 12. Assessing map robustness to the exclusion of sera.** Each serum group was removed and the map reoptimized. Arrows point to the position of each variant in the map shown in Figure X, for color correspondence refer to this map. A small arrow length indicates similar variant positions and map robustness to the exclusion of the particular serum group. Triangles point to sera positioned outside the plotting area. Maps without **A** mRNA-1273/mRNA-1273, **B** ChAdOx-S1/ChAdOx-S1, **C** ChAdOx-S1/BNT162b2, **D** BNT162b2/BNT162b2, **E** Ancestral virus conv., **F** alpha/alpha+E484K conv., **G** beta conv., **H** delta conv., **I** BA.1 omicron conv., **J** BA.2 omicron conv., **K** BA.5 omicron conv., **L** CK.2.1.1 conv..

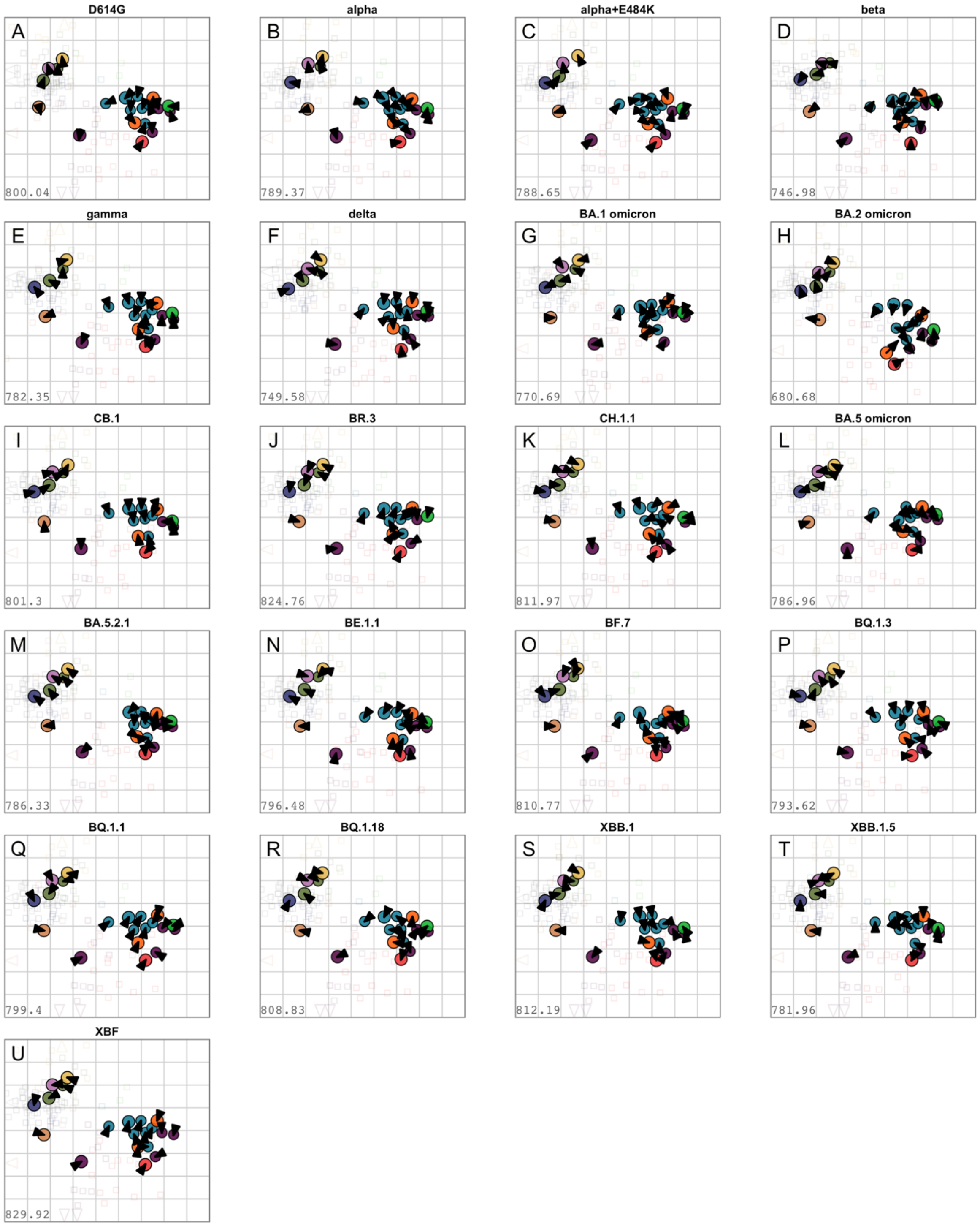


**Supplementary Figure 13. Assessing map robustness to the exclusion of antigen variant.** Each antigen variant was removed and the map reoptimized. Arrows point to the position of each variant in the map shown in Figure 3, for color correspondence refer to this map. A small arrow length indicates similar variant positions and robustness to the exclusion of the particular antigen variant. Triangles point to sera positioned outside the plotting area. Maps without **A** D614G, **B** alpha, **C** alpha+E484K, **D** gamma, **E** beta, **F** delta, **G** BA.1 omicron, **H** BA.2 omicron, **I** CB.1, **J** BR.3, **K** CH.1.1, **L** BA.5 omicron, **M** BA.5.2.1, **N** BE.1.1, **O** BF.7**, P** BQ.1.3, **Q** BQ.1.1, **R** BQ.1.18, **S** XBB.1, **T** XBB.1.5, **U** XBF.

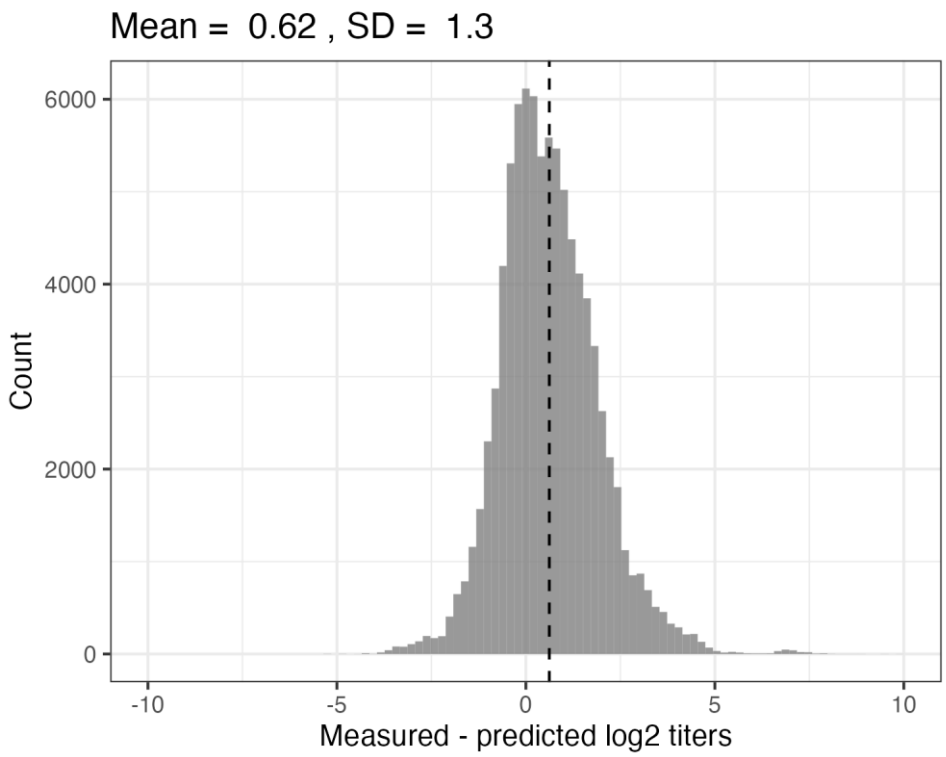


**Supplementary Figure 14. Map cross-validation residual titers.** 1000 repeats with 1000 optimization runs each were performed with only 90% of measured titers used for map construction by artificially masking 10% of measurements. The missing $\log_2$ titers were predicted by subtracting the Euclidean map distance for each serum-antigen pair from the maximum $\log_2$ titer of the specific serum. The difference between predicted and detectable measured titers on the $\log_2$ scale was calculated, the mean is indicated by the dashed line. The mean and mean-centered standard deviation are given.

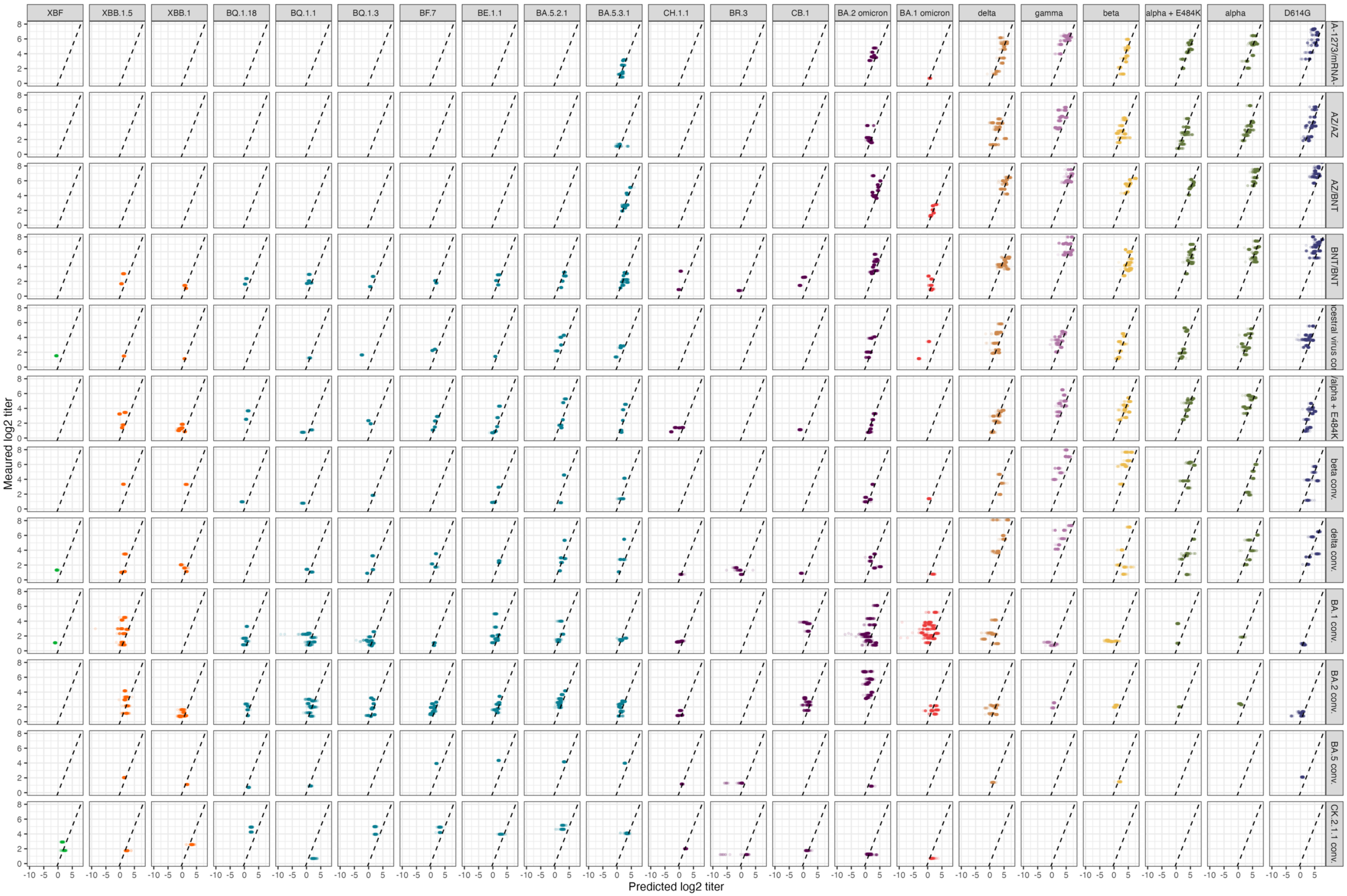


***Supplementary Figure 15: Map cross-validation predicted vs. measured titers.*** *Figure caption on next page.*

**Supplementary Figure 15. Map cross-validation predicted vs. measured titers.** 1000 repeats with 1000 optimization runs each were performed with only 90% of measured titers used for map construction by artificially masking 10% of measurements. The missing $\log_2$ titers were predicted by subtracting the Euclidean map distance for each serum-antigen pair from the maximum $\log_2$ titer of the specific serum. The detectable measured over predicted log2 titers are shown per serum group and antigen variant. The lower x-axis limit has been set to -10 for plotting purposes, very few residuals in the omicron convalescent groups were larger than that due to inaccurate positioning of sera.

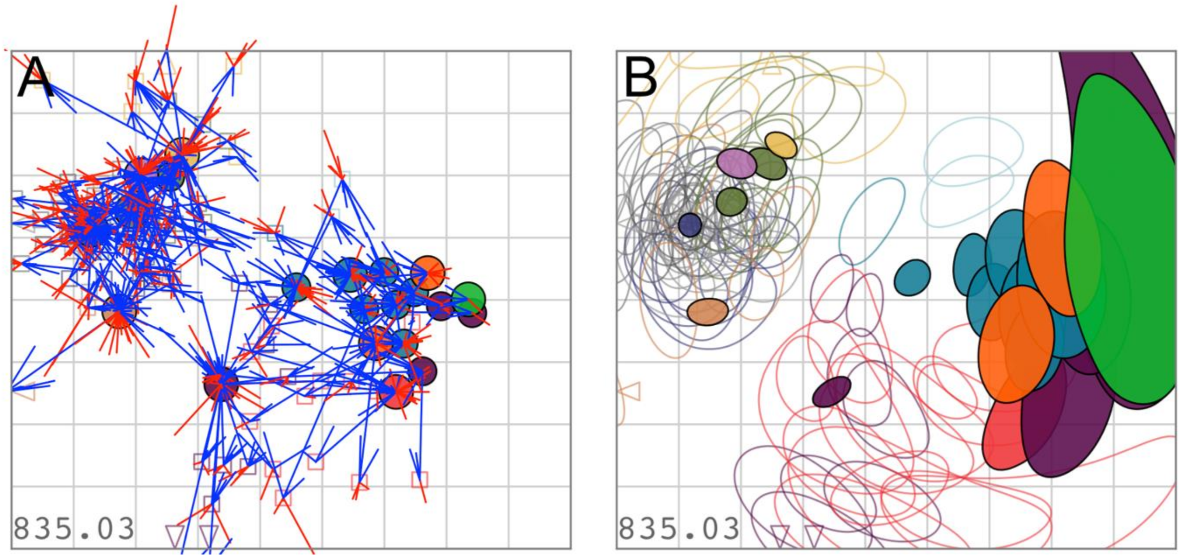


**Supplementary Figure 16. Titer error lines and map triangulation. A** Error lines for each serum and antigen are shown in blue in case of larger map distance than target distance and red in case of smaller map distance than target distance. The length of each error bar indicates the magnitude of mismatch. Blue error lines point towards the variant-serum pair that has a smaller target distance, red error lines point away from the variant-serum pair. **B** Constant force loci (Triangulation blobs) show the area for each serum and variant in which the item can move without increasing map stress by more than one unit. Filled shapes show variant Triangulation blobs, open shapes sera. Colors correspond to the map shown in Figure X.

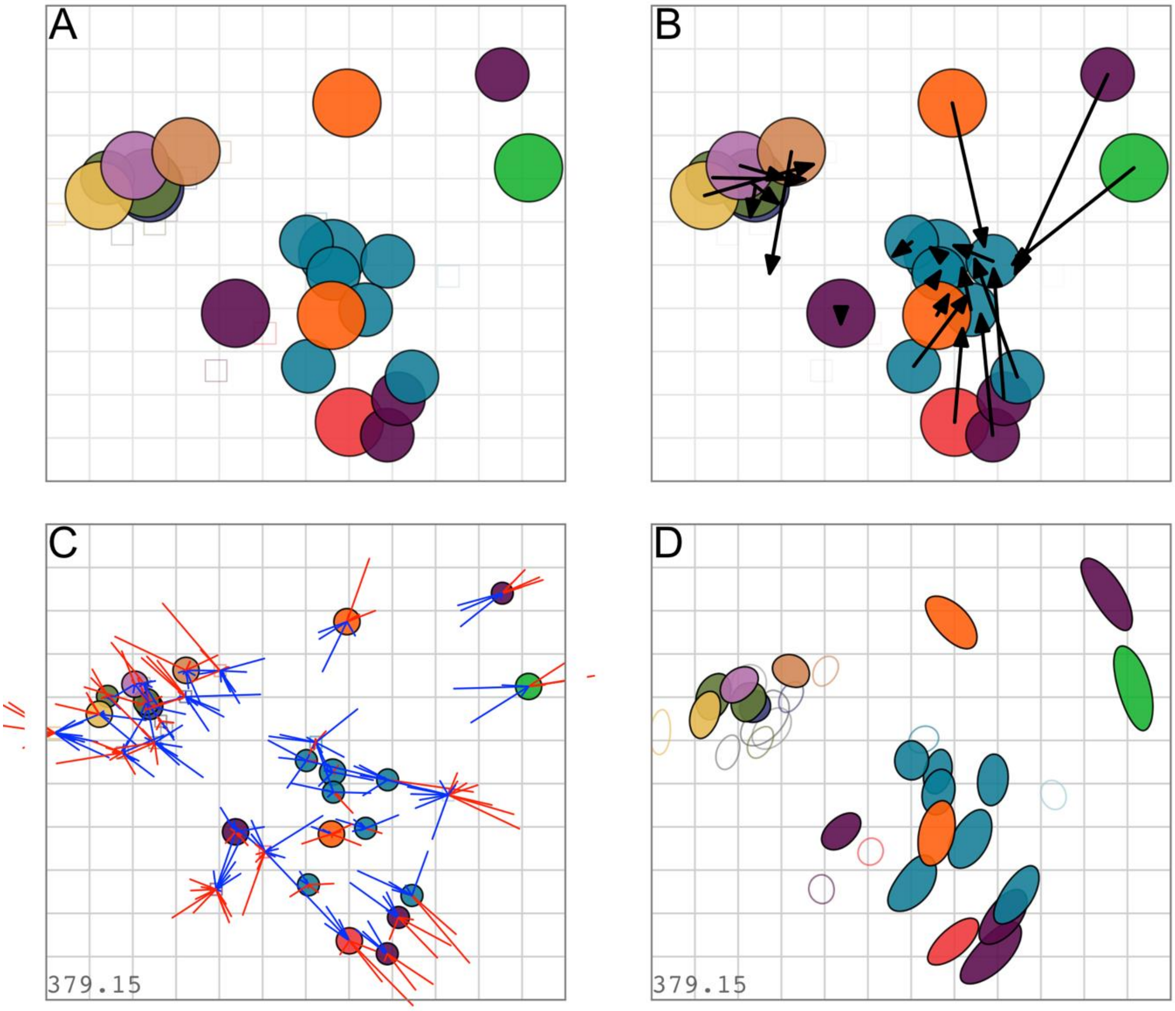


**Supplementary Figure 17. Map from geometric mean titers GMT. A** GMTs of each serum groups were used for map construction. The colors correspond to the colors used in Figure X. **B** Map in A with arrows pointing towards the variants' positions in Figure 3. **C** Error lines connecting GMT sample and variant. Blue lines indicate a larger map than target distance, red lines indicate a smaller map than target distance. **D** Constant force foci (Triangulation blobs) of variants and GMT samples. The marked area corresponds to the area an item can occupy without increasing the map stress by more than 1 unit. Filled shapes show variant Triangulation blobs, open shapes sera.

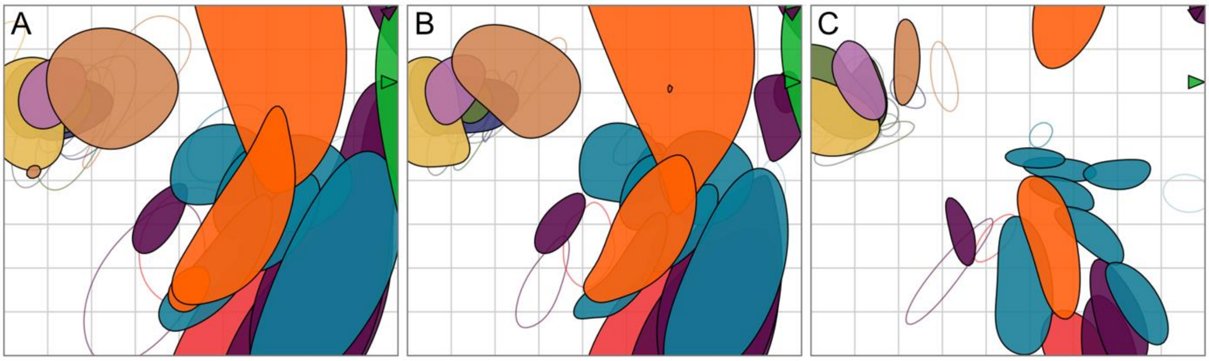


**Supplementary Figure 18. Assessing GMT map robustness to measurement uncertainty by bootstrapping.** 500 bootstrap repeats were performed with 1000 optimizations per repeat. In each repeat, different weights are assigned to each part of the titer table. The weights are drawn randomly from a Dirichlet distribution. Different weights were added to **A** titers and antigen reactivity, **B** only titers, and **C** only antigen reactivity. The colored regions mark 68% (one standard deviation) of the positional variation for each variant (filled shapes) and sera (open shapes). The colors correspond to the colors used in Figure 3.

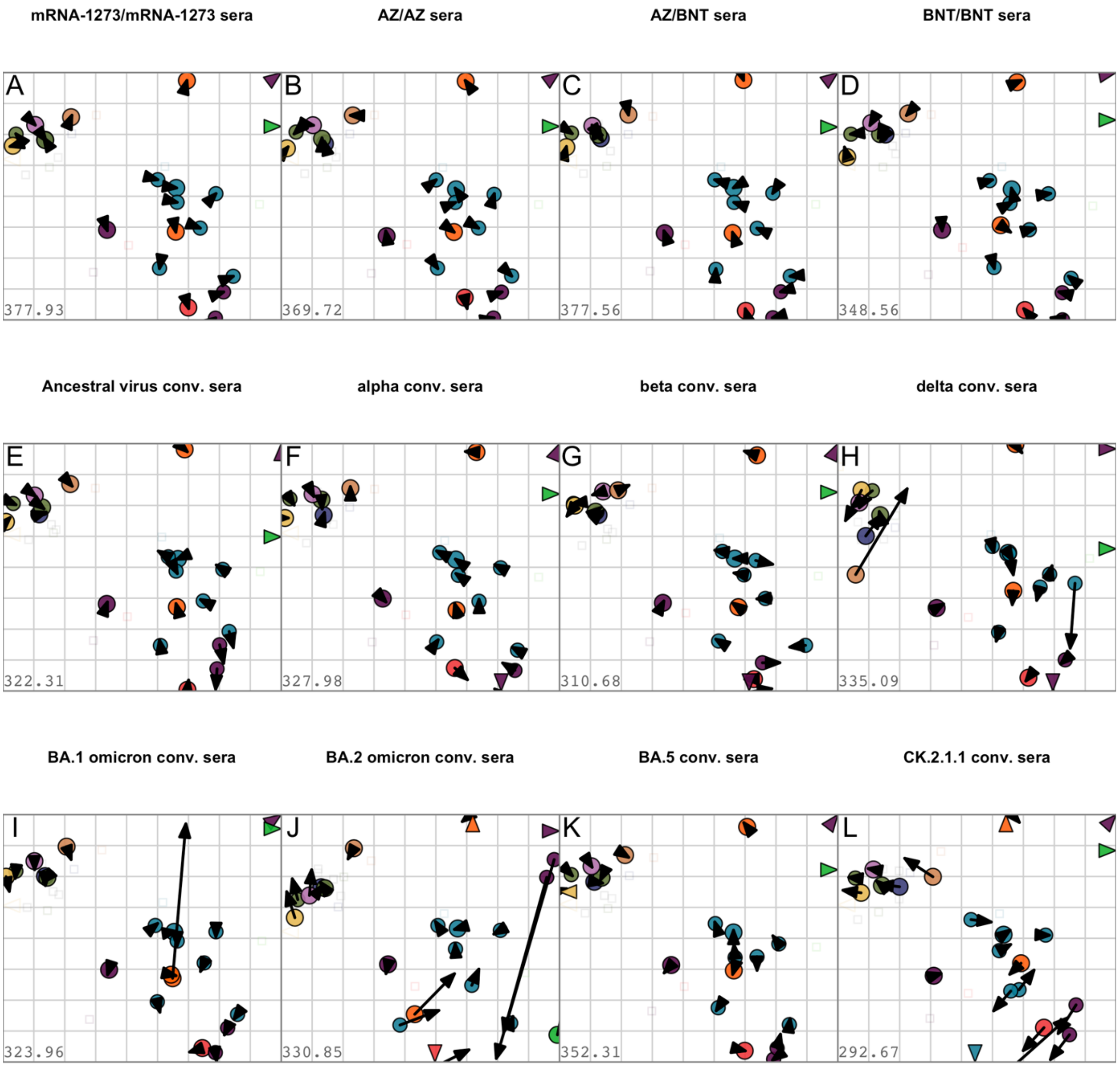


**Supplementary Figure 19. Assessing GMT map robustness to the exclusion of sera.** Each serum group was removed and the map re-optimized with optimization 1000 iterations. Arrows point to the position of each variant in the gmt map shown in Supplementary Figure 17A, for color correspondence refer to the map in the main manuscript. A small arrow length indicates similar variant positions and map robustness to the exclusion of the particular serum group. Maps without **A** mRNA-1273/mRNA-1273, **B** ChAdOx-S1/ChAdOx-S1, **C** ChAdOx-S1/BNT162b2, **D** BNT162b2/BNT162b2, **E** Ancestral virus conv., **F** alpha/alpha+E484K conv., **G** beta conv., **H** delta conv., **I** BA.1 omicron conv., **J** BA.2 omicron conv., **K** BA.5 omicron conv., **L** CK.2.1.1 conv..

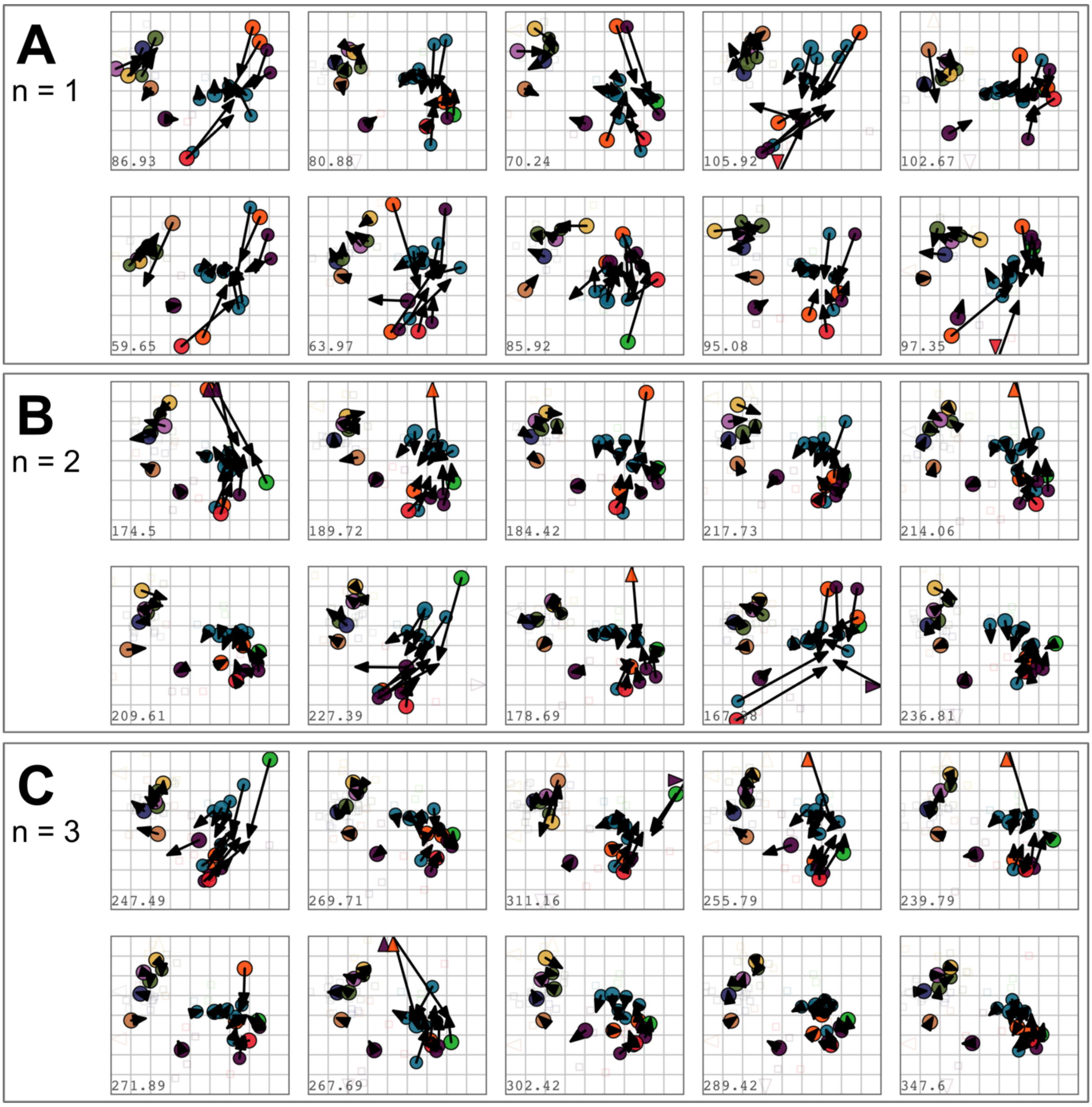


**Supplementary Figure 20. Assessing map robustness to sample size per serum group.** Ten Maps were created with a randomly drawn subset of samples with different sample sizes. 1000 optimizations were performed per map with a dilution step size of 0 and the minimum column basis set to "none". **A** n=1, **B** n=2, **C** n=3 samples were randomly drawn per serum group.

A.

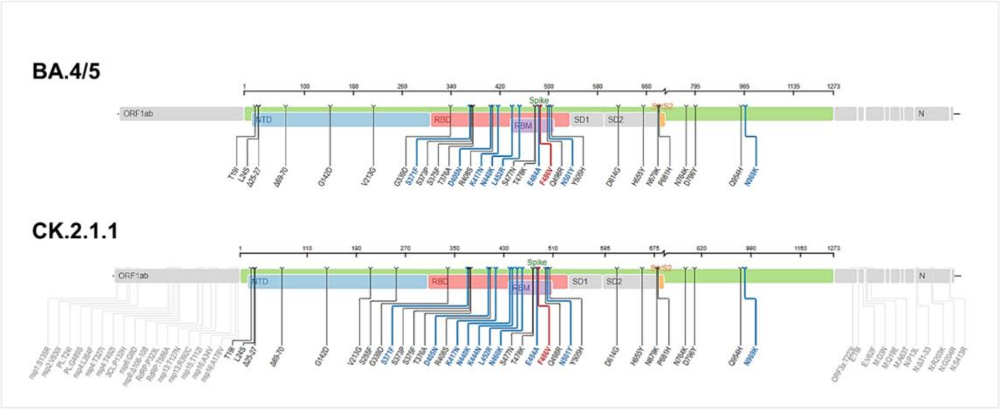


B.

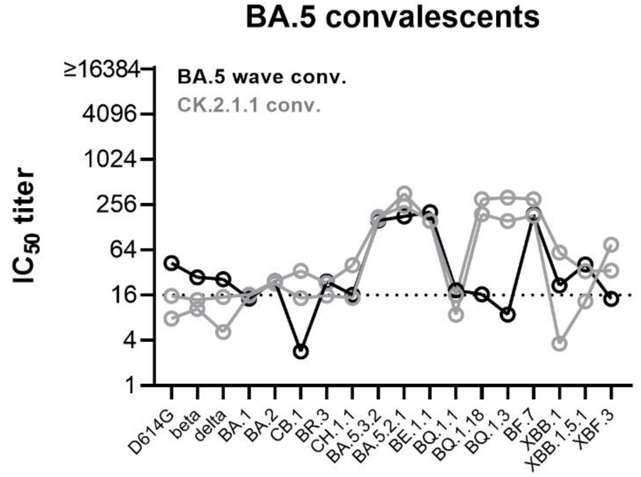


**Supplementary Figure 21. Neutralization profile of BA.5 omicron convalescent individuals.** (A) Spike mutations in BA.5 compared to CK.2.1.1, generated using https://covdb.stanford.edu/sierra/sars2/by-sequences/. (B) Neutralizing antibody titers against D614G, beta, delta, BA.1, BA.2, CB.1, BR.3, CH.1.1, BA.5.3.2, BA.5.2.1, BE.1.1, BF.7, BQ.1.3, BQ.1.8, BQ.1.1, XBB.1, XBB.1.5.1, and XBF.3 variants for one BA.5 first exposure (black) and two CK.2.1.1 first exposure sera (gray) are shown. Values for each patient are connected by a line. Titers below 16 were treated as negative (dotted line).

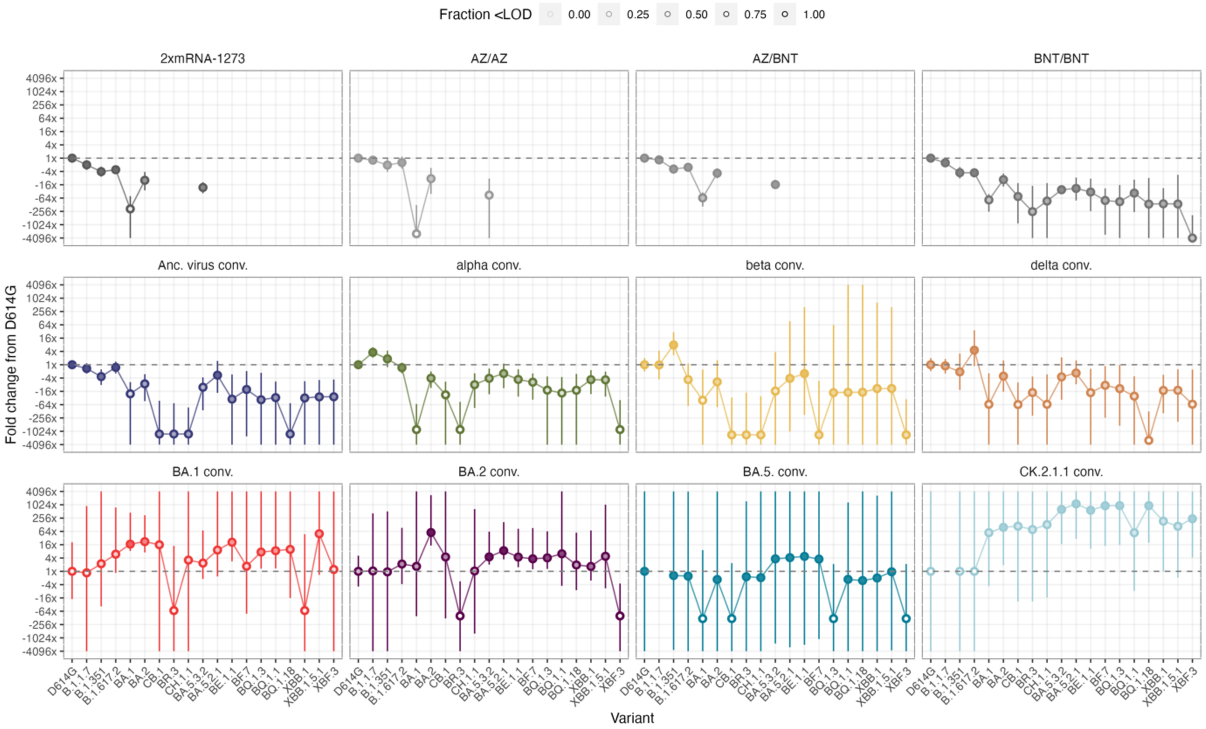


**Supplementary Figure 22. Titer fold changes from D614G for map serum groups.** Mean fold changes from D614G and 95% CI were calculated for each map serum group using the titertools R package[1], where below threshold values are interpolated using a Bayesian approach. The variants on the x-axis are ordered by decreasing fold change from D614G in the BNT/BNT group. The whiteness of each point corresponds to the fraction of titers <LOD, increasing with the number of samples <LOD. For fully colored circles, all samples had detectable titers against the respective variant. Part of the data used for calculation of fold changes have been published previously in [2-4].

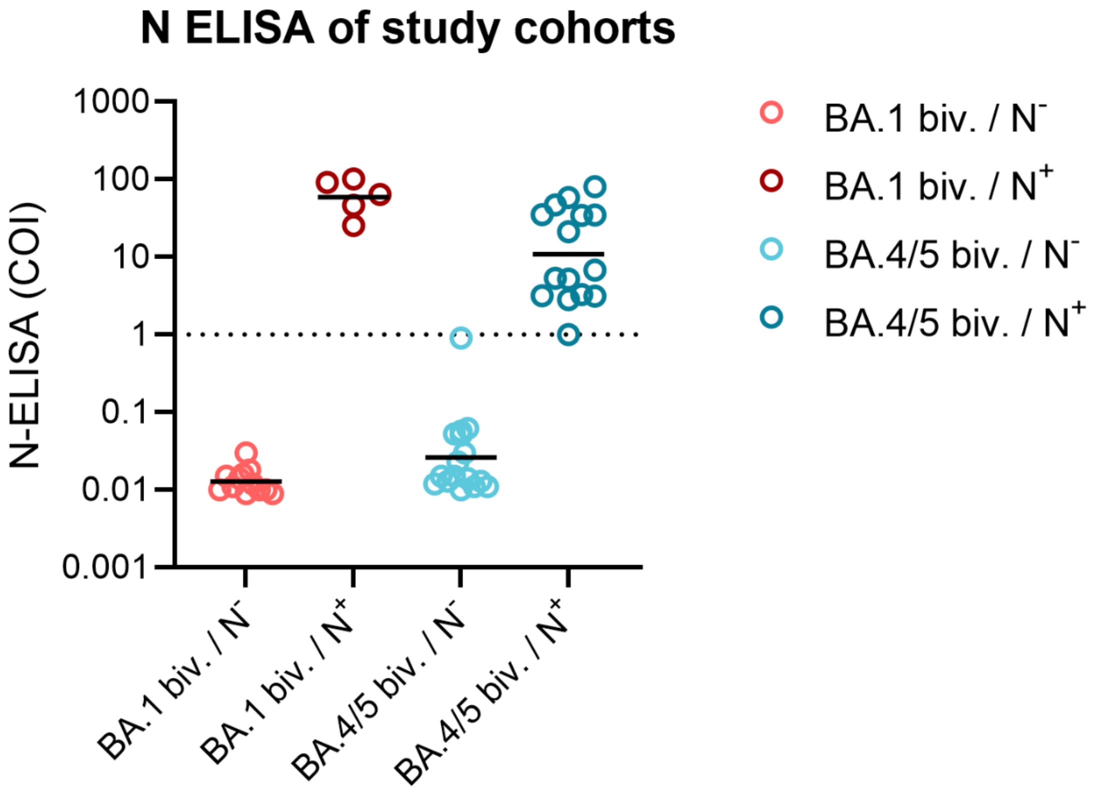


**Supplementary Figure 23. Grouping of cohorts according to anti-nucleocapsid antibodies.** Plasma samples were tested for anti-nucleocapsid (N) antibodies using the Elecsys Anti-N assay by Roche. Cut-off index (COI) ≥ 1 were treated as positive and samples were grouped accordingly into N negative (without infection history) and N positive (with previous SARS-CoV-2 infection) individuals. Shown are individual values (n=13 for BA.1 biv./$N^-$, n=5 for BA.1 biv./$N^+$, n=17 for BA.4/5 biv./$N^-$, n=16 for BA.4/5 biv./$N^+$) and mean.

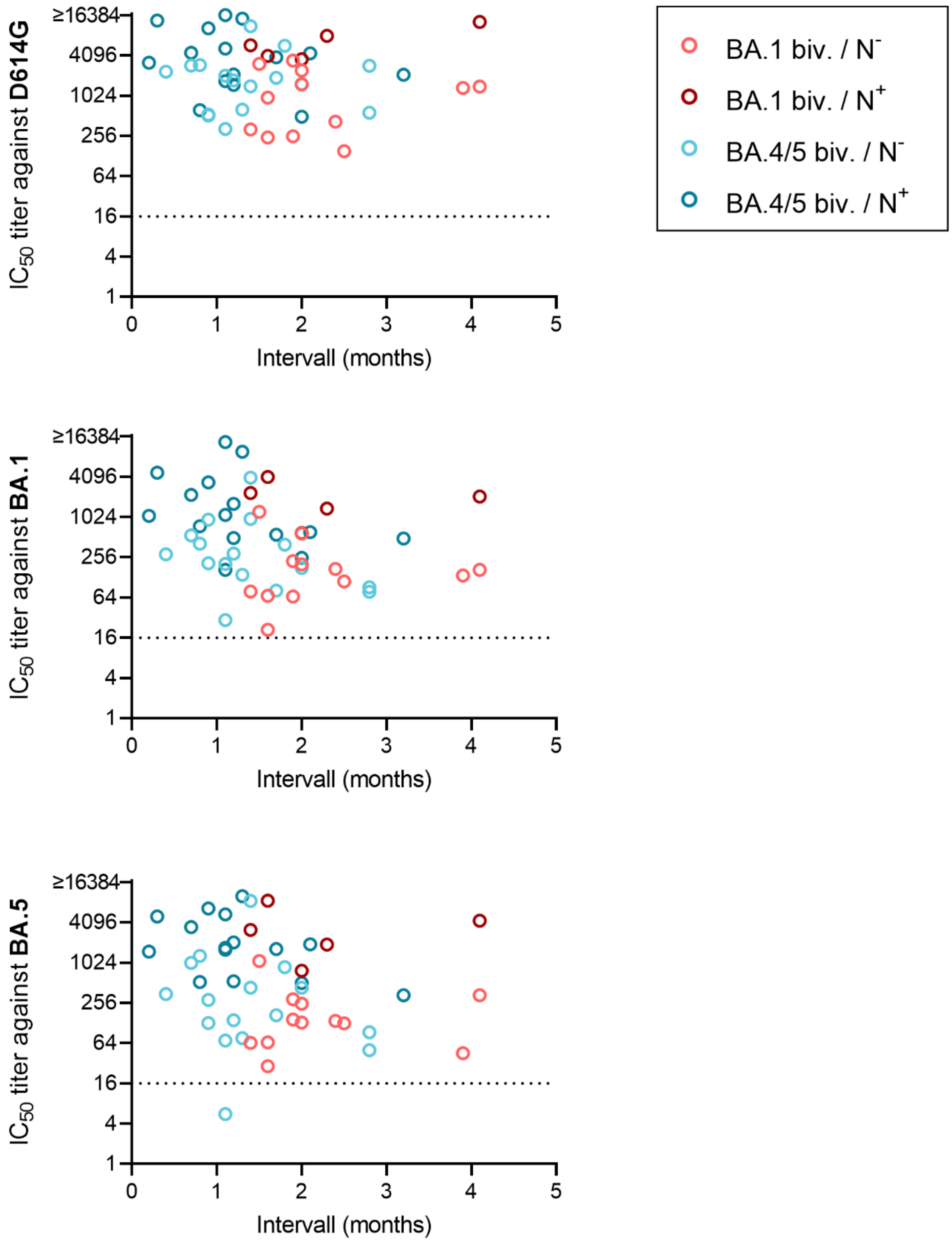


**Supplementary Figure 24. Neutralizing antibodies relative to interval between bivalent booster and blood collection.** Titers of neutralizing antibodies against D614G, BA.1 and BA.5 were blotted against the interval between bivalent booster and blood collection. $IC_{50}$ titers below 16 were treated as negative (indicated by dotted lines).

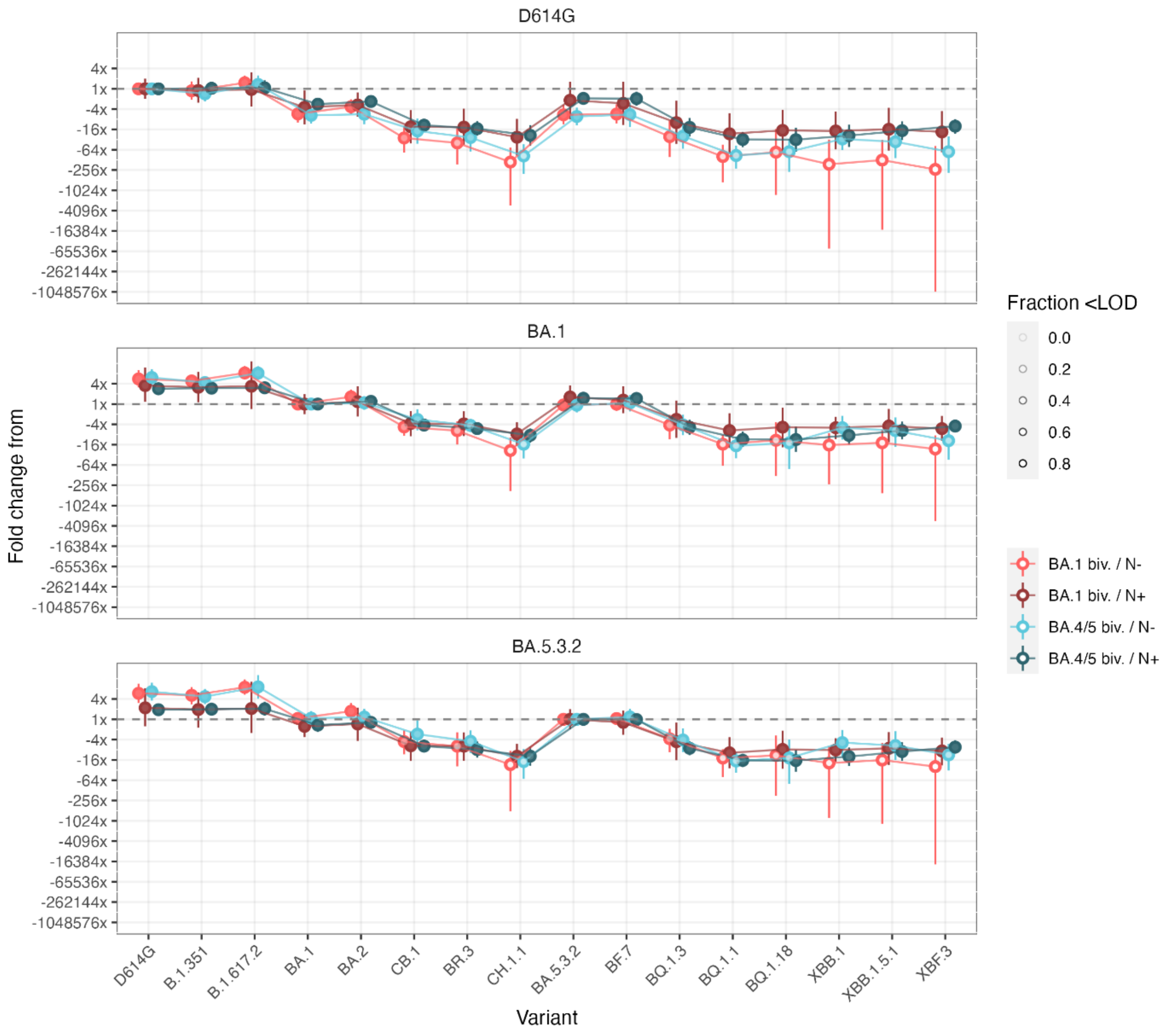


**Supplementary Figure 25. Titer fold changes for bivalent vaccine groups.** Mean fold changes from D614G (top graph), BA.1 (middle graph) or BA.5.3.2 (lower graph) to variants indicated on the x axis and 95% CI were calculated for each bivalent vaccine group using the titertools R package,[1] where below threshold values are interpolated using a Bayesian approach. The whiteness of each point corresponds to the fraction of titers <LOD, increasing with the number of samples <LOD. For fully colored circles, all samples had detectable titers against the respective variant.

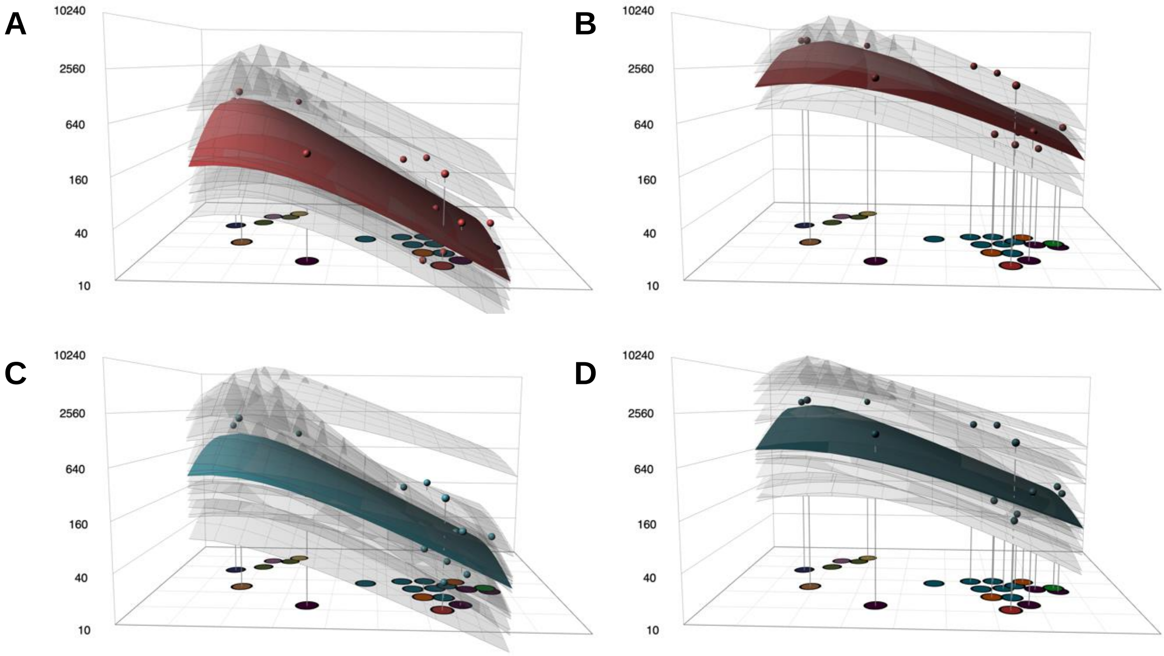


**Supplementary Figure 26. Individual antibody landscapes.** Antibody landscapes were fit for each serum in the different bivalent vaccine cohorts. Individual landscapes are shown as grey transparent surfaces, the GMT landscape of each vaccine cohort is shown as fully opaque surface, the GMTs against each variant are represented by small circles above the corresponding variant. The antibody landscapes were fit as described in the methods section. **A** BA.1 biv. / $N^-$, **B** BA.1 biv. / $N^+$, **C** BA.4/5 biv. / $N^-$, **D** BA.4/5 biv. / $N^+$.

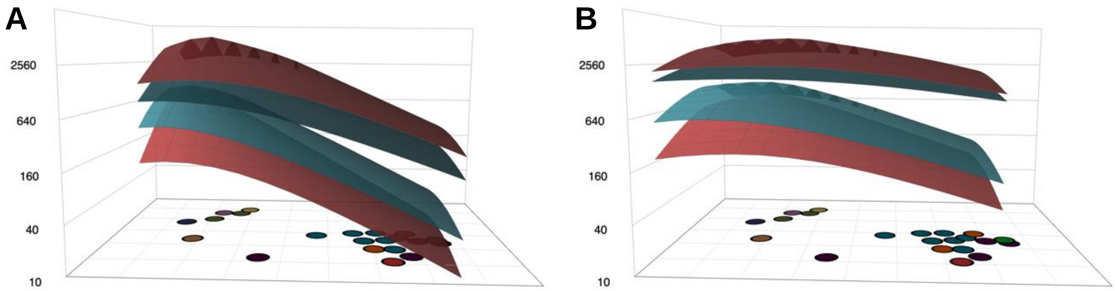


**Supplementary Figure 27. Antibody landscapes fit to a subset of variants.** GMT antibody landscapes were fit for the different bivalent vaccine cohorts to **A** pre-omicron, early-omicron and late-omicron variants or **B** to pre-omicron and early omicron variants (excluding CB.1, BR.3, CH.1.1, BA.5.2.1, BE.1.1, BF.7, BQ.1.3, BQ.1.1, BQ.1.18, XBB.1, XBB.1.5, XBF). The antibody landscapes were fit as described in the methods section and the colors encode the bivalent vaccine cohort (red: BA.1 biv. / $N^-$, dark red: BA.1 biv. / $N^+$, turquoise: BA.4/5 biv. / $N^-$, dark turquoise: BA.4/5 biv. / $N^+$).

### Supplementary Tables

#### Supplementary Table 1. Patient characteristics

| | | | | | Anti-nucleocapsid ELISA | | SARS-CoV-2 infection | |
|---|---|---|---|---|---|---|---|---|
| **Study cohort** | **Number of Participants** | **Mean age [years] ± SD** | **% female (number)** | **Mean interval between bivalent booster and blood collection [months] ± SD** | **Number positive** | **Mean COI ± SD** | **% with known infection history (number)** | **Variant#** |
| BA.1 biv./$N^-$ | 12 | 60.3 ± 10.9 | 50 (6) | 2.2 ± 0.8 | 0 | - | 0 | - |
| BA.1 biv./$N^+$ | 5 | 71.8 ± 10.8 | 60.0 (3) | 2.3 ± 1.0 | 5 | 65.9 ± 28.2 | 0 (0) | - |
| BA.4/5 biv./$N^-$ | 16 | 66.4 ± 15.3 | 68.8 (11) | 1.4 ± 0.7 | 0 | - | - | - |
| BA.4/5 biv./$N^+$ | 15 | 62.6 ± 21.3 | 60.0 (9) | 1.3 ±0.7 | 15 | 22.9 ± 24.0 | 37.5 (4) | alpha (n=1)<br>BA.2 omicron (n=3) |

n: number; COI: cut-off index, COI ≥ 1 was considered positive as specified by the manufacturer
#For two of the omicron BA.2 convalescent individuals infecting virus variant was determined by sequencing or melting curve analysis; for one BA.2 and the alpha convalescent individual infecting virus variant was assumed based on time point of infection.

**Supplementary Table 2. Virus variants used.**

| Isolate ID* | Pango lineage§ | Category | GISAID ID |
|---|---|---|---|
| B86.2 | B.1.177 (D614G; ancestral) | pre-omicron | EPI_ISL_3305837 |
| C63.1 | B.1.1.7 | pre-omicron | EPI_ISL_3277382 |
| C79.2 | B.1.1.7 (E484K) | pre-omicron | EPI_ISL_3277383 |
| C24.1 | B.1.351 | pre-omicron | EPI_ISL_17528983 |
| D94 | P.1.1 | pre-omicron | EPI_ISL_2095177 |
| D27 | B.1.617.2 | pre-omicron | EPI_ISL_2290769 |
| E16.1 | BA.1 | BA.1 omicron | EPI_ISL_17528984 |
| E65.1 | BA.2 | BA.2 omicron | EPI_ISL_12486408 |
| F14.2 | CB.1 | BA.2 omicron (BA.2.75) | EPI_ISL_16679179 |
| F69.1 | BR.3 | BA.2 omicron (BA.2.75) | EPI_ISL_17076114 |
| G19.2 | CH.1.1 | BA.2 omicron (BA.2.75) | EPI_ISL_16744347 |
| E73.1 | BA.5.3.2 | BA.5 omicron | EPI_ISL_13666092 |
| F05.2 | BA.5.2.1 | BA.5 omicron | EPI_ISL_17528948 |
| F11.2 | BE.1.1 | BA.5 omicron | EPI_ISL_17528949 |
| F96.2 | BF.7 | BA.5 omicron | EPI_ISL_16744346 |
| F26.1 | BQ.1.3 | BA.5 omicron | EPI_ISL_17528950 |
| F74.1 | BQ.1.1 | BA.5 omicron | EPI_ISL_17076149 |
| F80 | BQ.1.18 | BA.5 omicron | EPI_ISL_17077092 |
| G22.3 | XBB.1 | recombinant | EPI_ISL_17076150 |
| G37.2 | XBB.1.5.1 | recombinant | EPI_ISL_17077093 |
| G45.1 | XBF.3 | recombinant | EPI_ISL_17324524 |

*Internal name of isolate; §determined using UShER (https://genome.ucsc.edu/cgi-bin/hgPhyloPlace) on 24.03.2023

**Supplementary Table 3. Overview of map sera titrated against each variant.***

| Variant | mR1273/ mR1273 | BNT/BNT | AZ/BNT | AZ/AZ | Anc. virus conv. | alpha/alpha+ E484K conv. | beta conv. | delta conv. | BA.1 conv. | BA.2 conv. | BA.5 conv. | CK.2.1.1 conv. |
|---|---|---|---|---|---|---|---|---|---|---|---|---|
| D614G | 10 | 11 | 10 | 10 | 10 | 9 | 6 | 5 | 14 | 10 | 1 | 2 |
| B.1.1.7 | 10 | 11 | 10 | 10 | 10 | 9 | 6 | 5 | 14 | 10 | - | - |
| B.1.1.7+E484K | 10 | 11 | 10 | 10 | 10 | 9 | 6 | 5 | 14 | 10 | - | - |
| B.1.351 | 10 | 11 | 10 | 10 | 10 | 9 | 6 | 5 | 14 | 10 | 1 | 2 |
| P.1.1 | 10 | 11 | 10 | 10 | 10 | 9 | 6 | 5 | 14 | 10 | - | - |
| B.1.617.2 | 10 | 11 | 10 | 10 | 10 | 9 | 6 | 5 | 14 | 10 | 1 | 2 |
| BA.1 | 10 | 11 | 10 | 10 | 10 | 9 | 6 | 5 | 14 | 10 | 1 | 2 |
| BA.2 | 10 | 11 | 10 | 10 | 10 | 9 | 6 | 5 | 14 | 10 | 1 | 2 |
| CB.1 | - | 6 | - | - | 5 | 9 | 3 | 5 | 13 | 10 | 1 | 2 |
| BR.3 | - | 6 | - | - | 5 | 9 | 3 | 5 | 13 | 10 | 1 | 2 |
| CH.1.1 | - | 6 | - | - | 5 | 9 | 3 | 5 | 13 | 10 | 1 | 2 |
| BA.5.3.2 | 10 | 11 | 10 | 10 | 10 | 9 | 6 | 5 | 14 | 10 | 1 | 2 |
| BA.5.2.1 | - | 6 | - | - | 5 | 9 | 3 | 5 | 13 | 10 | 1 | 2 |
| BE.1.1 | - | 6 | - | - | 5 | 9 | 3 | 5 | 13 | 10 | 1 | 2 |
| BF.7 | - | 6 | - | - | 5 | 9 | 3 | 5 | 13 | 10 | 1 | 2 |
| BQ.1.3 | - | 6 | - | - | 5 | 9 | 3 | 5 | 13 | 10 | 1 | 2 |
| BQ.1.1 | - | 6 | - | - | 5 | 9 | 3 | 5 | 13 | 10 | 1 | 2 |
| BQ.1.18 | - | 6 | - | - | 5 | 9 | 3 | 5 | 13 | 10 | 1 | 2 |
| XBB.1 | - | 6 | - | - | 5 | 9 | 3 | 5 | 13 | 10 | 1 | 2 |
| XBB.1.5 | - | 6 | - | - | 5 | 9 | 3 | 5 | 13 | 10 | 1 | 2 |
| XBF.3 | - | 6 | - | - | 5 | 9 | 3 | 5 | 13 | 10 | 1 | 2 |

*A previous map was extended to include more recent variants of concern and BA.5 and CK.2.1.1 conv. (= convalescent) sera. Not all sera that were used to construct the previous map were titrated against the additional variants due to low volume.
